# Deciphering the genetic architecture of atrial fibrillation offers insights into disease prediction, pathophysiology and downstream sequelae

**DOI:** 10.1101/2023.07.20.23292938

**Authors:** Shuai Yuan, Yuying Li, Lijuan Wang, Fengzhe Xu, Jie Chen, Michael G Levin, Ying Xiong, Benjamin F. Voight, Scott M Damrauer, Dipender Gill, Stephen Burgess, Agneta Åkesson, Karl Michaëlsson, Xue Li, Xia Shen, Susanna C. Larsson

## Abstract

**Aims:** The study aimed to discover novel genetic loci for atrial fibrillation (AF), explore the shared genetic etiologies between AF and other cardiovascular and cardiometabolic traits, and uncover AF pathogenesis using Mendelian randomization analysis.

**Methods and results:** We conducted a genome-wide association study meta-analysis including 109,787 AF cases and 1,165,920 controls of European ancestry and identified 215 loci, among which 91 were novel. We performed Genomic Structural Equation Modeling analysis between AF and four cardiovascular comorbidities (coronary artery disease, ischemic stroke, heart failure, and vneous thromboembolism) and found 189 loci shared across these diseases as well as a universal genetic locus shared by atherosclerotic outcomes (i.e., rs1537373 near *CDKN2B*). Three genetic loci (rs10740129 near *JMJD1C*, rs2370982 near *NRXN3*, and rs9931494 near *FTO*) were associated with AF and cardiometabolic traits. A polygenic risk score derived from this genome-wide meta-analysis was associated with AF risk (odds ratio 2.36, 95% confidence interval 2.31-2.41 per standard deviation increase) in the UK biobank. This score, combined with age, sex, and basic clinical features, predicted AF risk (AUC 0.784, 95% CI 0.781-0.787) in Europeans. Phenome-wide association analysis of the polygenic risk score identified many AF-related comorbidities of the circulatory, endocrine, and respiratory systems. Phenome-wide and multi-omic Mendelian randomization analyses identified associations of blood lipids and pressure, diabetes, insomnia, obesity, short sleep, and smoking, 27 blood proteins, one gut microbe (*genus.Catenibacterium*), and 11 blood metabolites with risk to AF.

**Conclusions:** This genome-wide association study and trans-omic Mendelian randomization analysis provides insights into disease risk prediction, pathophysiology and downstream sequelae.

## Introduction

Atrial fibrillation (AF) is a common arrhythmia, characterized by disorganized atrial depolarizations, which can lead to symptoms including palpitations and decreased exercise capacity, as well as more serious complications such as heart failure, stroke, and death. With an aging global population, AF has become an epidemic and important health issue with increasing incidence and prevalence, ^1^ particularly in North America and Europe ^2^. The Global Burden of Disease 2019 Study estimated that approximately 59.7 million individuals live with AF, which is associated with 8.4 million disability-adjusted life years worldwide. ^3^ Hence, there is an urgent need to elucidate the pathological basis of AF to improve prevention and treatment.

Alongside environmental factors, the contribution of genetic factors to the pathogenesis of AF has been increasingly recognized. Several genome-wide association studies (GWASs) have been conducted to disentangle the genetic architecture of AF and uncovered over 100 loci involved in AF development. ^4–8^ Despite this, these GWASs explain a small portion of the estimated heritability. This gap between observed and estimated heritability suggests that additional AF-associated variants remain to be discovered. A GWAS with a larger sample size may empower the identification of rarer variants and variants with smaller effects. Additionally, by identifying genetic predictors of AF, it will be possible to prioritize the clinical development of therapeutic targets.^9^

Randomized controlled trials, observational, and genetic studies have implicated several modifiable risk factors in the pathogenesis of AF, including hypertension, obesity, smoking, poor sleep, etc.^10–15^ Mendelian randomization (MR) analysis is an epidemiological approach that can reinforce causal inference by using genetic variants as an instrumental variable for the exposure under three key assumptions. ^16^ The current availability of GWAS data on a broad spectrum of measurements, including circulating proteins, gut microbiota, and metabolites, has enabled efficient approaches to exploring the etiology of AF using MR design. These associations, including for circulating proteins that can reflect therapeutic targets, ^15^ may benefit strategy formulation for disease prevention and drug development.

To further facilitate the understanding of the genetic etiology of AF and elucidate the underlying genetic architecture, we conducted an updated GWAS meta-analysis involving up to 1.3 million individuals. Moreover, we investigated shared genetic signals between AF and cardiovascular comorbidities and cardiometabolic traits. We also examined the risk prediction ability of AF polygenic risk score and AF’s causal consequences using a polygenic risk score phenome-wide association design. Finally, based on this updated GWAS meta-analysis, we conducted omics-MR analyses to illuminate the pathogenesis of AF.

## Methods

### Study design and participants

**Figure 1** shows the study design. We performed a GWAS meta-analysis and downstream analyses to understand genetic and molecular architectures of AF. This GWAS meta-analysis included data from three sources (a previous meta-analysis of 6 studies, ^6^ the FinnGen study R8,^17^ and the SIMPLER cohorts [https://www.simpler4health.se/]). The descriptions of included studies (definition, genotyping array, and imputation) are shown in **Supplementary Methods** and **Table S1**. Ethical committees had approved all studies, and participants had signed informed consent forms. We then performed subsequent analyses to prioritize gene candidates, reveal the genetic etiologies linking AF, cardiovascular comorbidities, and cardiometabolic traits, examine the utility of genetic information in AF risk prediction, and explore the risk factors for AF from different perspectives using omics data.

**Figure 1.**
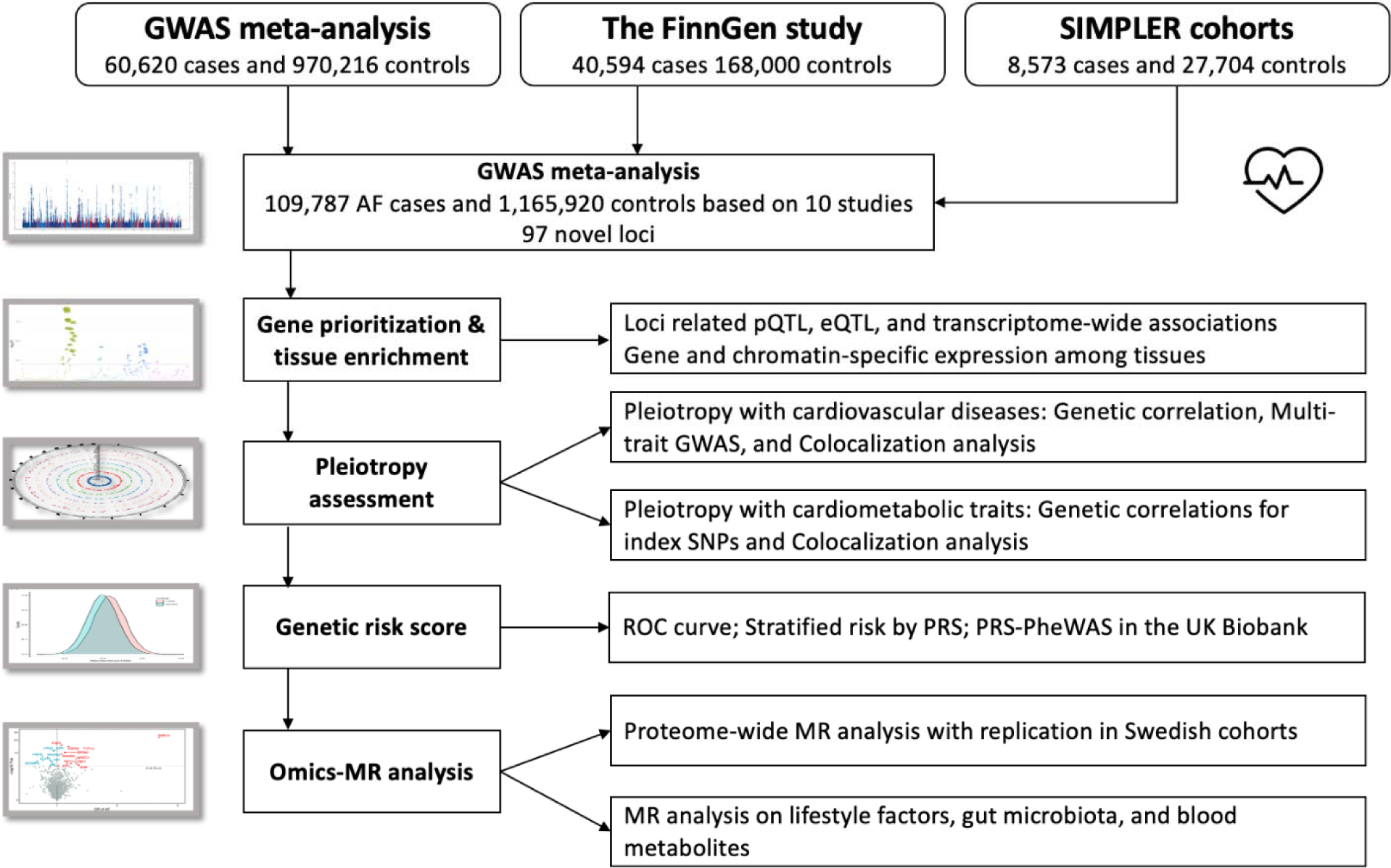
Study design overview. Abbreviations: GWAS, genome-wide association study; MR, Mendelian randomization; PRS-PheWAS, polygenetic risk score-phenome-wide association analysis; ROC, operating characteristic curve; SIMPLER, Swedish Infrastructure for Medical Population-based Life-course and Environmental Research.

### Genome-wide association analysis

In the GWAS meta-analysis, we included three data sources (Nielsen et al GWAS, FinnGen R8, and SIMPLER) with 109,787 AF cases and 1,165,920 controls. The quality control was conducted at the marker and sample levels for each included study (**Supplementary methods)**. In brief, each dataset underwent initial quality control, imputation, post-imputation quality control, and association tests with at least age (birth year), sex, and principal components as covariates. We meta-analyzed these data using METAL with the fixed-effect inverse-variance-weighted method.^18^ Genomic inflation factor (λ _GC_) was calculated for the GWAS meta-analysis. To assess any residual confounding due to population stratification, we calculated the linkage disequilibrium score regression (LDSC) intercept using SNP (single nucleotide polymorphism) LD scores calculated in the HapMap3 CEU population ^19^. Independent significant genomic risk loci were defined as: 1) *P*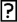<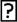5×10^-8^; 2) window 500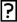kb; 3) linkage disequilibrium *r*^2^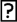=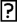0.6 and *r* ^2^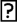=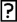0.1 (a common setting in clumping independent loci in GWAS^20^), and the pruning process was conducted using FUMA with the 1000 Genomes Phase 3 European reference panel.^21^

### Gene prioritization and tissue-specific enrichment

We prioritized genes located within 10 kb of the lead variant for each locus using three methods: 1) coding variants. Gene type is based on gene biotype obtained from BioMart (Ensembl 85);^22^ 2) eQTL mapping. The lead variants at each risk locus were mapped to genes using eQTL data from GTEx v.8 of whole blood, blood vessels (artery aorta, artery coronary, and artery tibial), heart (heart atrial appendage and left ventricle), and lung; and 3) transcriptome-wide association study (TWAS). TWAS in whole blood, blood vessels (artery aorta, artery coronary, and artery tibial), heart (heart atrial appendage and left ventricle), and lung was based on the application of S-MultiXcan integrating with GTEx v8 gene expression and splicing data (**Supplementary methods**).^23, 24^ We utilized LDSC-SEG ^25^ to examine the enrichment of disease heritability by integrating our GWAS-meta-analysis summary statistics with gene expression ^26^ and chromatin^27^ datasets. To account for multiple testing, we employed false discovery rate (FDR) correction individually for each dataset with a significance threshold of FDR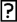<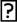0.05. We also used FUMA to obtain differentially expressed gene sets for each of the 53 tissue types based on the Genotype-Tissue Expression (GTEx) project dataset.^21^

### Pleiotropy with cardiovascular diseases

Cross-trait LDSC and high-definition likelihood method(HDL)^28^ were performed to estimate genetic correlations of AF with related cardiovascular diseases, including heart failure (HF),^29^ coronary artery disease (CAD), ^30^ ischemic stroke (ISSTROKE), ^31^ and venous thromboembolism (VTE)^32^ with data from corresponding GWASs. LDSC and HDL employ GWAS summary data to estimate SNP heritability (the proportion of phenotypic variance explained by measured SNPs) and genetic correlation between polygenic traits, while considering sample overlap and linkage disequilibrium information. We then used Genomic Structural Equation Modeling (Genomic-SEM)^33^ to obtain the joint genetic architecture of the above traits. The Genomic-SEM technique can estimate genetic correlation, measure heritability, evaluate interdependence among traits, and accommodate complete sample overlap.^33^ Its versatility lies in the ability to employ equations to model proposed connections between observed traits and latent variables. To determine SNP-level effects, the genetic covariance and sampling covariance matrices are expanded to incorporate SNPs, which are then subjected to individual regression based on the parameters specified by each structural model. We used a common factor model. Model specifications can be found in the **Supplementary methods**. The analysis was implemented using the GenomicSEM package in R. ^33^ To explore whether the loci identified in Genomic SEM share a genetic etiology, we used HyPrColoc,^34^ a recently developed Bayesian algorithm designed to simultaneously and efficiently evaluate for colocalization across multiple traits. We first assessed for colocalization across AF, HF, CAD, ISSTROKE, and VTE. We conducted sensitivity analyses where we implemented modifications to the regional and alignment thresholds, raising the values from 0.6 to 0.9, and adjusted the colocalization prior, experimenting with values of 0.02, 0.01, and 0.005.

### Pleiotropy with cardiometabolic traits

We first calculated the genetic correlations of AF with seven cardiometabolic traits. To assess the pleiotropic effects of AF-associated SNPs, we obtained the associations of lead SNPs in 215 loci with seven cardiometabolic traits, including BMI,^35^ waist-to-hip ratio, ^35^ low- and high-density lipoprotein cholesterol,^36^ triglycerides,^36^ systolic blood pressure,^37^ and type 2 diabetes^38^. Colocalization analysis was performed for the associations between AF-associated loci and cardiometabolic traits.^39^

### Polygenic risk score (PRS) regression

We selected independent SNPs associated with AF at the *P*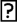<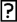5×10^-8^ in the GWAS meta-analysis and without linkage disequilibrium (*r*^2^ < 0.001) to construct PRS. To reduce the risk of bias from sample overlap, the weights for SNPs in the PRS were obtained from the GWAS meta-analysis of FinnGen and SIMPLER studies after excluding Nielsen et al GWAS that contains the UK Biobank. The weighted PRS was created by summing the number of AF-liability-increasing alleles for each SNP weighted by the log-transformed odds ratio of AF and then adding this weighted score for all used SNPs. We estimated the associations of the PRS in tertiles with AF (36,886 prevalent and incident cases out of 385,917 unrelated White British individuals in the UK Biobank study) using logistic regression with adjustment for age^2^, sex, assessment center, and the first 10 principal components. For PRS in a continuous manner, we used the area under the receiver operating characteristic curve (AUC) to compare the discriminatory ability of the PRS relative to PRS plus nongenetic factors, like age, sex, and cardiometabolic risk markers (i.e., body mass index, high- and low-density lipoprotein cholesterol, triglycerides, and systolic blood pressure).

### PRS-phenome-wide association study (PRS-PheWAS)

We performed a PRS-PheWAS in the UK Biobank to explore the comorbidities associated with AF. The PRS-PheWAS was based on 1,060 phenotypes with number of cases > 200. The phenotypes were defined by the PheCODE schema based on ICD-9 and ICD-10 codes.^40^ The associations were estimated by a logistic regression model with adjustment for age^2^, sex, assessment center, and the first 10 principal components. The Bonferroni method was used to correct for multiple testing (*P* < 4.7×10^-5^). Details of the PRS-PheWAS can be found elsewhere. ^41^

### Multiple omics-wide Mendelian randomization analysis

Based on the GWAS meta-analysis data, we performed MR analysis to estimate the associations of 26 modifiable factors, 2,076 blood proteins, 211 gut microbiotas, and 352 annotated metabolites and metabolite ratios with AF risk. Detailed introduction to the MR design is provided in **Supplementary Methods**. The GWAS data sources are described in **Table S2.** We selected genetic variants associated with the exposures of interest at the significance level of *P*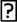<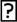5×10^-8^. Independent SNPs were used as an instrumental variable after pruning these SNPs at *r*^2^ < 0.01 to minimize the effect of collinearity of selected SNPs in linkage disequilibrium. For MR analysis of blood proteins, we used index cis-SNPs associated with the levels of plasma proteins at *P* < 5×10^-8^ as instrumental variables. Cis-SNPs were defined as SNPs within 1Mb from the gene encoding the protein and linkage disequilibrium was estimated based on 1000 Genomes European panel. We calculated *F* statistics^42^ to assess the strength of instrumental variables and found that all F statistics were > 10. For traits with SNPs ≤ 3, we used the inverse variance weighted method under the fixed effect model to estimate the MR association with AF. Otherwise, the inverse variance weighted method under the multiplicative random effects model was used. For traits with three or more SNPs, the weighted median and MR-Egger regression methods were applied to test the consistency of the results. Cochran’s Q test examined heterogeneity among SNPs’ estimates. The MR-Egger intercept test was used to evaluate the potential existence of horizontal pleiotropy. Colocalization analysis (**Supplementary methods**) based on cis gene region was used for blood proteins to rule out the possibility that the association was caused by linkage disequilibrium.

We conducted a traditional epidemiological association analysis (the prospective cohort analysis) in the SIMPLER cohorts to replicate certain MR associations for blood proteins, measured using the Olink platform. Detailed information on proteomic profiling in these cohorts can be found elsewhere.^43^ For this analysis, we used multivariable adjusted Cox proportional hazards regression to estimate the associations between blood protein levels and future risk of AF in 10,796 individuals free of AF diagnosis at baseline (**Supplementary methods**).

## Results

### Genome-wide association analysis identified 91 novel loci

The GWAS meta-analysis included 109,787 AF cases and 1,165,920 controls, and ∼29.3 million sequence variants. The genomic inflation factor (λ_GC_) was 1.48, and the LDSC intercept was 1.09 (standard error = 0.03), suggesting that most of the inflation is due to AF polygenicity. The SNP heritability was estimated to be 11.2% (95% confidence interval (CI) 9.6%-12.8%) on the observed scale and 5.3% (95% CI 4.6%-6.1%) on the liability scale, assuming a disease prevalence of 0.51% ^44^. A total of 215 loci were identified at the conventional genome-wide significance threshold (*P*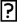<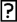5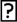×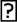10^-8^; **Table S3**), of which 91 loci are novel (based on prior AF signals found in the GWAS catalog, **Figure 2**). The strongest signal was observed for one locus near *PITX2*. Although 213 loci had directional consistency in effect size across studies, two (rs167479 near *RGL3* and rs2240128 near *DOT1L*) showed differences in effect size (*P*_HET_ <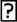0.05/215, **Table S3**). Most risk alleles conferred small-to-modest effect size with an odds ratio (OR) ranging from 1.03 to 1.23 per allele (**Figure S1**). Three lead SNPs had an OR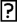>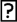1.3, including rs532342679 near *NEDD1* (novel loci), rs190065070 near *EMC10*, and rs147301839 near *GCOM1* (**Figure S1**). Of note, these are rare SNPs. The identification may be majorly due to an increased sample size instead of a genomic inflation since these SNP-AF associations were consistent across studies.

**Figure 2.**
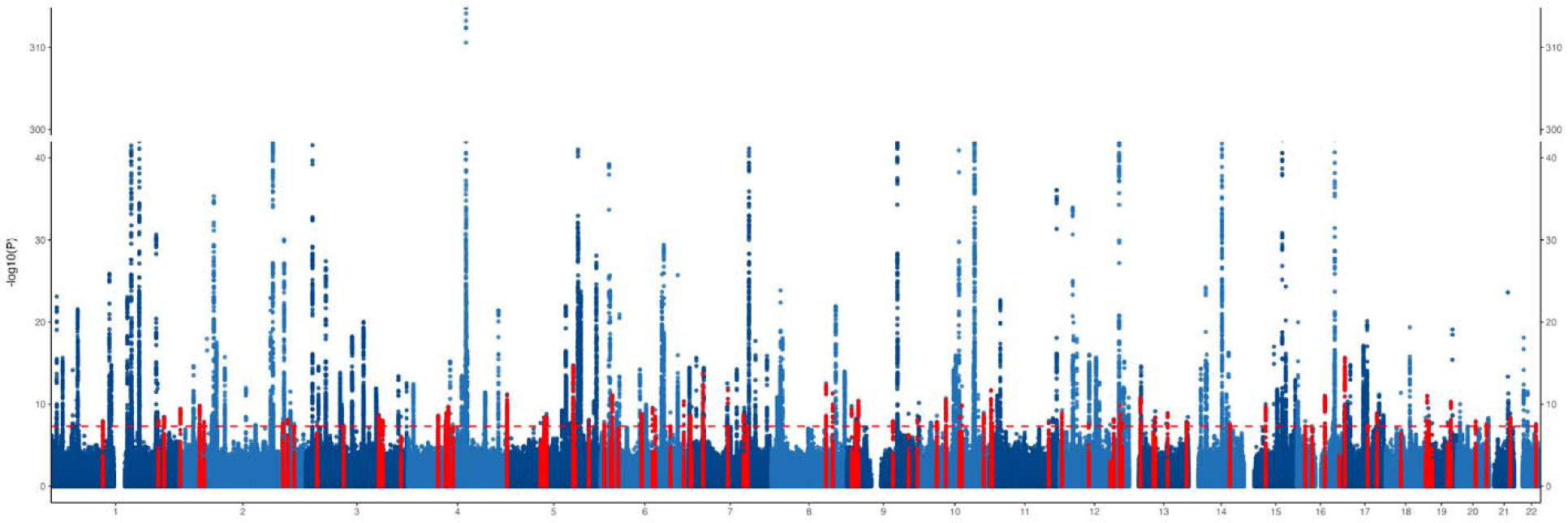
Manhattan plot of the results from atrial fibrillation GWAS meta-analysis. Each point represents a genetic variant. Genetic variants against the log-transformed *P* value of the associations with AF in the GWAS meta-analysis. Genetic variants in red represent variant located +/−500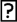kb of a novel genome-wide significant locus.

### In silico functional analyses prioritized loci

Result summary of loci prioritization is presented in **Table S4**. In coding variants annotation in BioMart (Ensembl 85), we found 31 loci with function of protein coding (**Table S4**). Seventy-eight loci were significantly expressed in selected tissues (**Table S5**). TWAS identified 153 loci with expression signals and 128 loci with splicing signals in the targeted tissues (**Table S6 and Table S7**). Nearby genes of loci identified were highly expressed in cardiovascular tissues (**Figure S2**), particularly in the heart atrial appendage and left ventricle (**Figure S3**).

### Pleiotropy with cardiovascular diseases

We detected moderate genetic correlations between AF and four other studied cardiovascular diseases (all *P* values < 7.13×10 ^-11^, **Figure S4**). In the Genomic SEM analysis, we identified 189 independent loci (**Table S8**). The number of loci with GWAS association at *P* < 5×10 ^-8^ ranged from 6 for HF to 103 for CAD, and 25 loci were defined as novel (**Figure 3A**). None of the loci were associated with all included outcomes. One locus (rs1537373 in *CDKN2B*) was associated with four outcomes, with the effect allele conferring consistent effects. Likewise, a locus near *LPA* conferred consistent effects on AF, CAD, and HF with strong colocalization support. AF shared 12 loci with CAD, 5 loci with HF, 4 loci with ISSTROKE, and 3 loci with VTE (**Figure 3B**). A total of 21 loci had moderate to high support of colocalization for AF associations and many showed pleiotropic effects on CAD and HF (**Figure 3C**).

**Figure 3.**
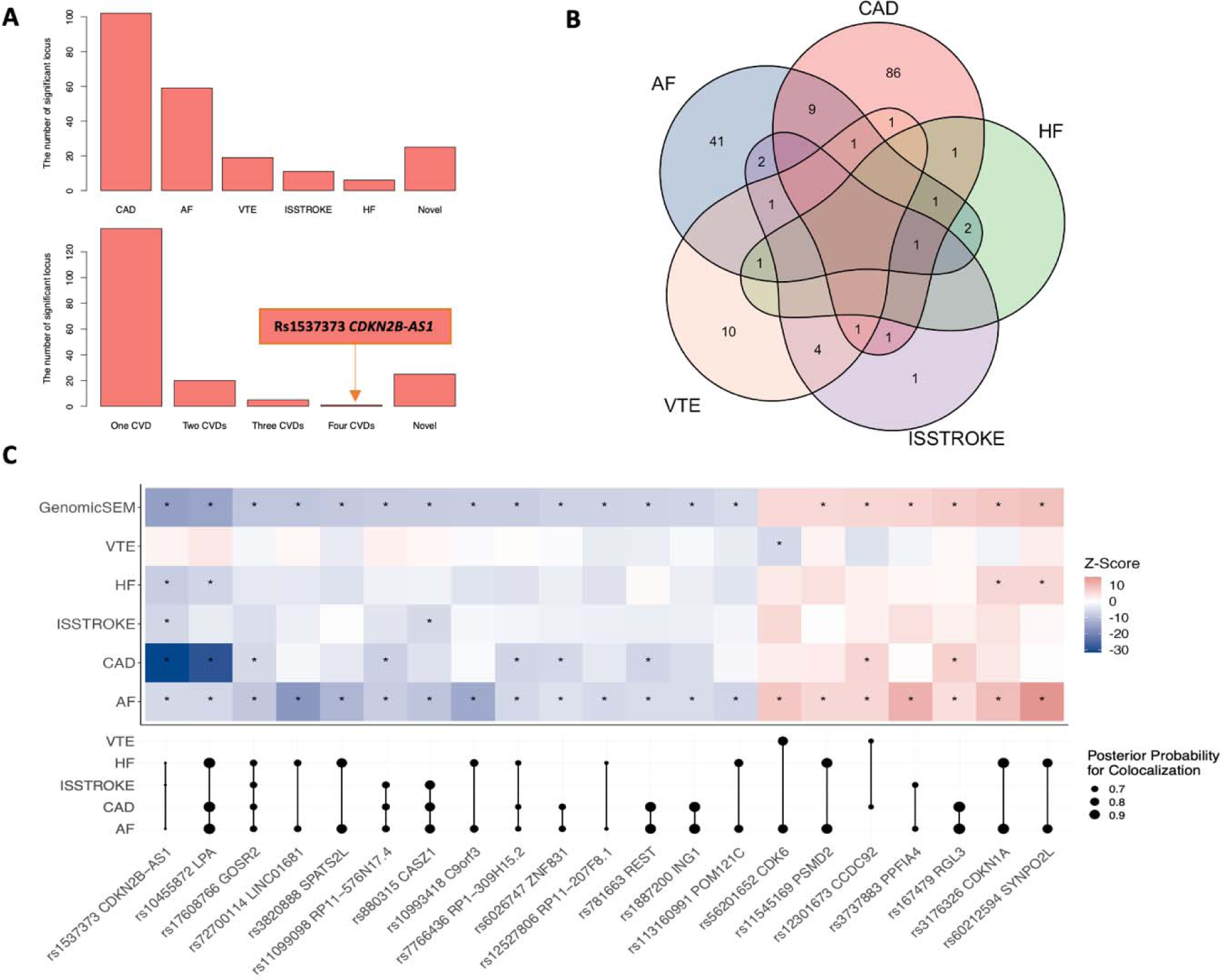
Genetic loci identified by Genomic SEM analysis and shared between atrial fibrillation and other cardiovascular diseases. Abbreviations: AF, atrial fibrillation; CAD, coronary artery disease; HF, heart failure; ISSTROKE, ischemic stroke; VTE, venous thromboembolism. **A**: number of loci associated with cardiovascular disease at the genome-wide significance level (upper) and number of loci associated with 0-4 cardiovascular diseases at the genome-wide significance level (lower). **B**: Venn plot of loci shared by studied cardiovascular diseases. **C**: The loci associated with AF and at least one other cardiovascular disease. Most these associations had moderate to strong colocalization support (the upper part shows the genetic associations and the lower part shows results of colocalization; the star sign means the *P* value < 5×10^-8^).

### Pleiotropy with cardiometabolic traits

We observed genetic correlations of AF with low-density lipoprotein cholesterol and waist-to-hip ratio (**Figure S5**). Among the 215 loci identified for AF, 56 loci were associated with at least one of the examined cardiometabolic traits at the *P*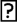<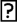5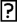×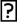10^-8^ (**Figure 4A**). We listed out 23 loci associated with AF and at least other two cardiometabolic traits at the *P*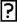<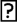5×10^-8^ (**Figure 4B**). Three loci (rs10740129 near *JMJD1C*, rs2370982 near *NRXN3*, and rs9931494 near *FTO*) were associated with at least six examined traits. Most these associations were supported by colocalization analysis (*PH4* > 0.8; **Figure 4B**).

**Figure 4.**
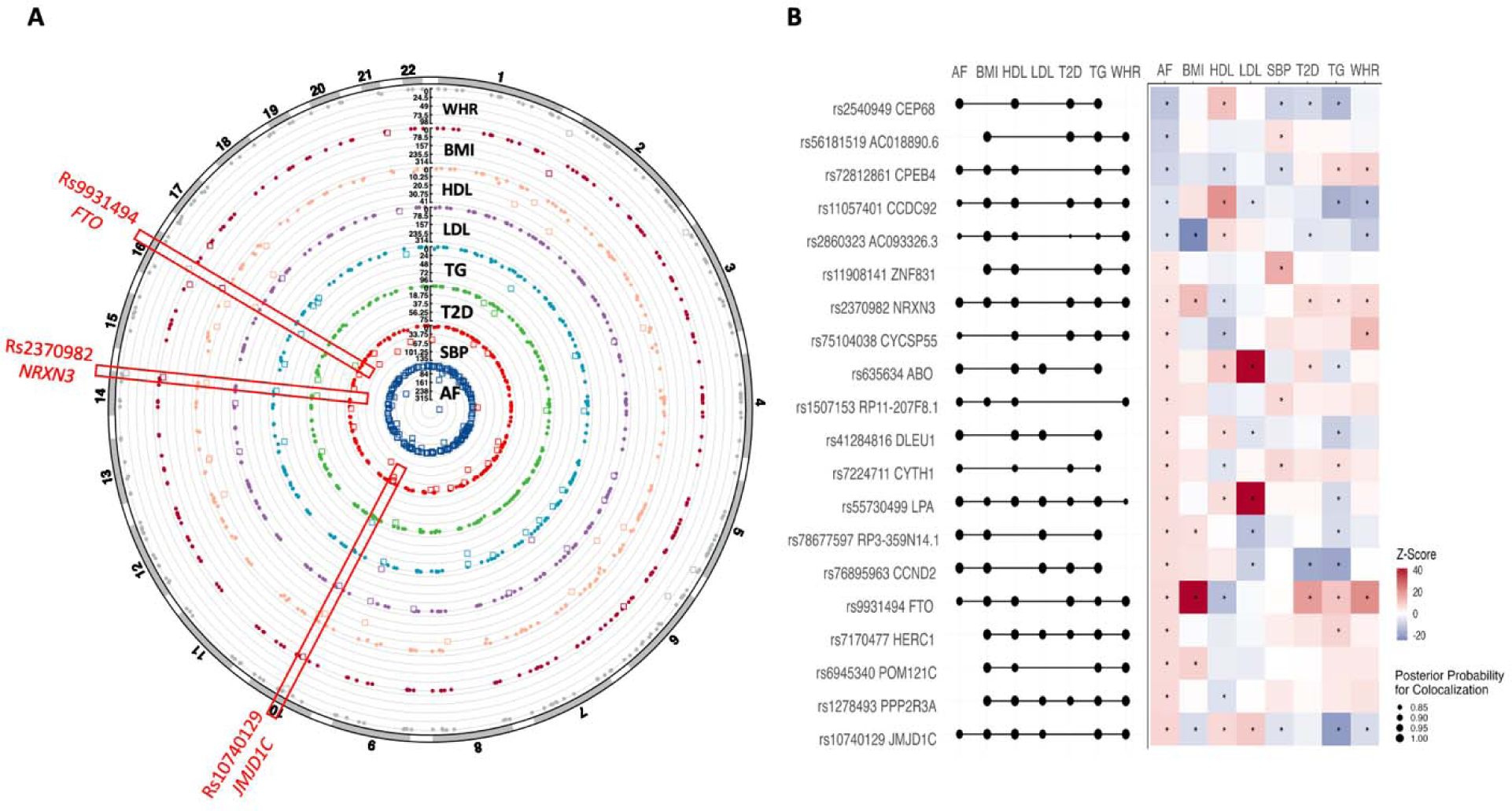
Pleiotropic effects of AF-associated loci with cardiometabolic traits. Abbreviations: AF, atrial fibrillation; BMI, body mass index; HDLC, high-density lipoprotein cholesterol; LDLC, low-density lipoprotein cholesterol; SBP, systolic blood pressure; T2D, type 2 diabetes; TG, triglycerides; WHR, waist-to-hip ratio. **A**: the circle plot of the associations of AF-associated loci with cardiometabolic traits. The associations with the *P* value < 5×10^-8^ were marked in square, otherwise in circle. **B**: AF-associated loci associated with at least one cardiometabolic trait and corresponding colocalization evidence (the right part shows the genetic associations and the left part shows results of colocalization; the star sign means the *P* value < 5×10^-8^).

### The polygenic risk score and PRS-PheWAS

Information on SNPs included in the PRS is shown in **Table S9**. The mean of standardized PRS was larger in AF cases compared to non-cases in the UK Biobank (**Figure 5A**). Comparing individuals with the lowest PRS score (tertile 1), the OR of AF was 1.33 (95% confidence interval (CI) 1.20-1.47) for those with the highest PRS score (tertile 3) (**Figure 5B**). When treating PRS in continuous, per standard deviation (SD) increase in AF-PRS, the OR of AF was 2.36 (95% CI 2.31-2.41; *P* < 0.001). The AUC for the model containing only the PRS (continuous) and nongenetic factors was 0.631 (95% CI 0.628-0.634) and 0.757 (95% CI 0.755-0.760), respectively (**Figure 5C**). The AUC increased for the model by adding PRS and nongenetic factors (0.784, 95% CI 0.781-0.787; **Figure 5C**). After corrections for multiple testing (*P* < 0.05/1060), 88 phenotypes were associated with the AF-PRS. Except for AF-related phenotypes (atrial fibrillation and flutter and cardiac arrhythmia), the AF-PRS was associated with high odds of heart failure, mitral valve disease, ischemic heart disease, hypertension, cardiomegaly, and other 49 diseases of the circulatory system, 8 endocrine/metabolic diseases, and 7 respiratory diseases (**Figure 5D and Table S10**).

**Figure 5.**
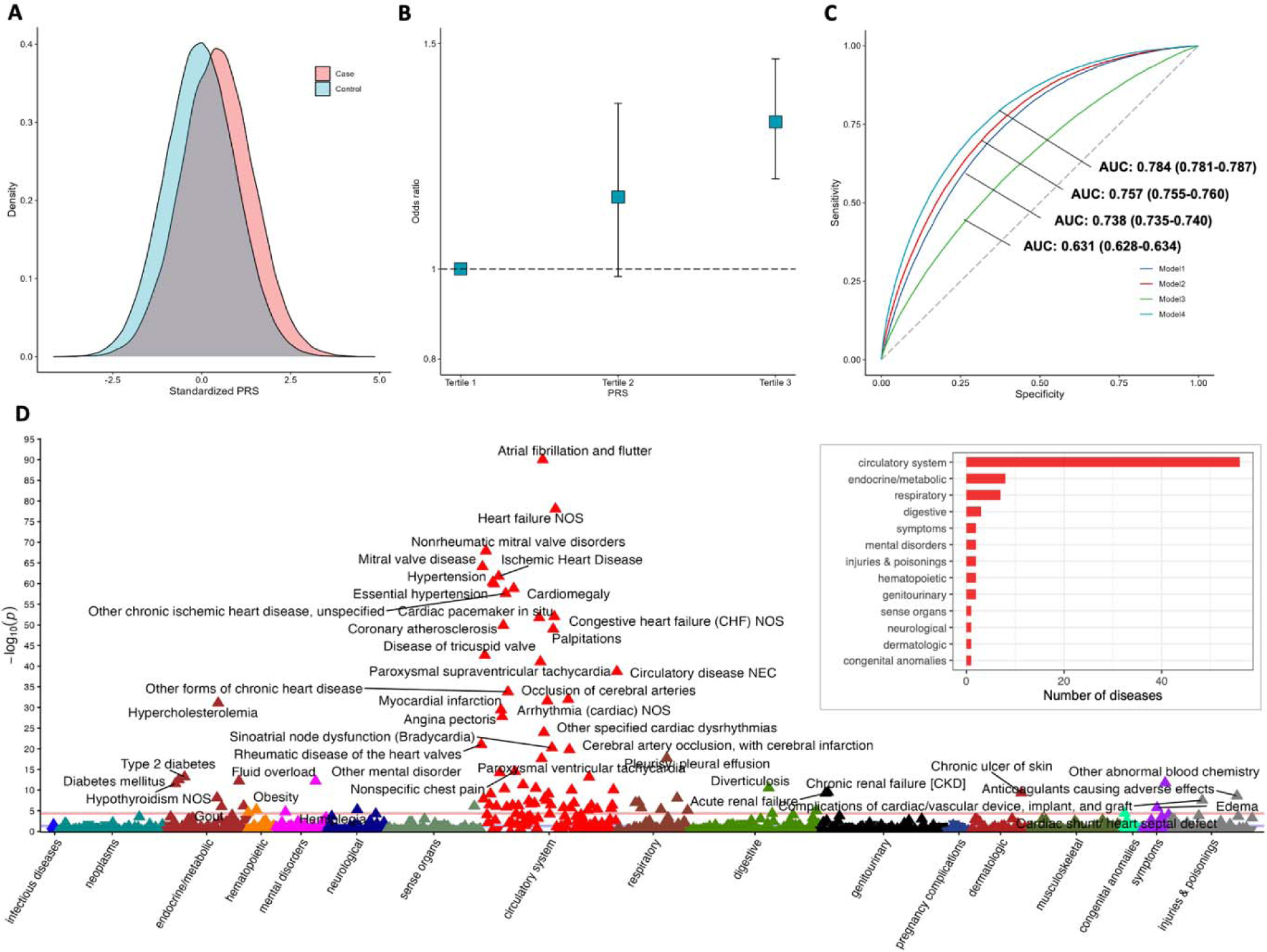
Associations of polygenic risk score (PRS) with the risk of atrial fibrillation and other phenotypes in the UK Biobank and the discriminatory ability of the PRS. **A**: distribution of PRS between AF cases and controls. **B**: odds ratio of AF by PRS tertiles. **C**: area under the receiver operating characteristic curve (AUC) to compare the discriminatory ability of the PRS relative to PRS plus nongenetic factors. Model 1 included age and sex; model 2 included age, gender, body mass index, high- and low-density lipoprotein cholesterol, triglycerides, and systolic blood pressure; model 3 included PRS; and model 4 included PRS plus all nongenetic factors above. **D**. results of PRS phenome-wide association analysis in the UK Biobank.

### Blood proteins and AF

After removing proteins without a genetic instrument in the AF GWAS meta-analysis dataset, the proteome-wide analysis included 1,887 proteins (**Table S11**). Genetically predicted levels of 27 circulating proteins were associated with AF risk after multiple testing corrections (*P* < 0.05/1887; **Figure 6A**). Per SD increase in genetically predicted protein levels, the OR of AF ranged from 0.67 (95% CI 0.57-0.79) for SCAMP3 (secretory carrier-associated membrane protein 3) to 2.69 (95% CI 2.22-3.27) for RAB1A (Ras-related protein Rab-1A) (**Figure 6B**). Among these proteins, two without summary-level data were excluded from colocalization analysis (**Table S12**). Four proteins had high support of colocalization with PH4 >0.8 (**Figure 6C**). Five protein-AF associations were tested in SIMPLER cohorts (**Table S13**). We replicated the association for ADM (adrenomedullin) protein measured by Olink CVD II panel in 10,913 participants free of baseline AF in an epidemiologic analysis of the SIMPLER cohorts. Per SD increase in ADM, the hazard ratio of incident AF was 1.28 (95% CI 1.17-1.40) in the model adjusted for batch, age, and sex (**Figure 6D**). The association remained in the analyses with further adjustment for lifestyle and the cardiometabolic risk markers (**Figure 6D, Table S13**).

**Figure 6.**
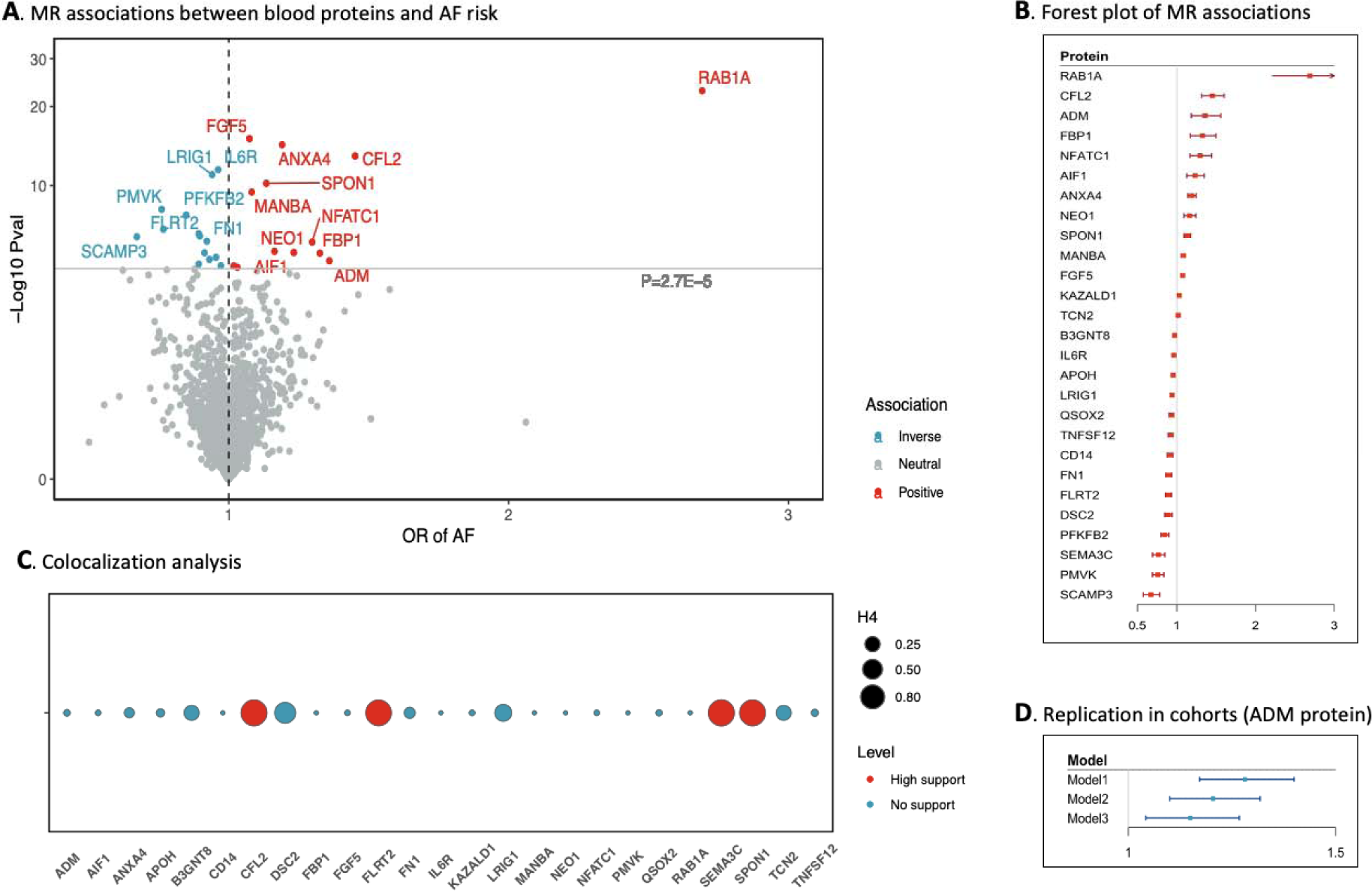
Proteome-wide Mendelian randomization analysis of atrial fibrillation and validation in SIMPLER cohorts. Abbreviations: AF, atrial fibrillation; MR, Mendelian randomization. **A**: 27 blood proteins associated with atrial fibrillation in MR analysis after Bonferroni correction. Names of these proteins are available in Table S11. **B**: Associations between 27 blood proteins and AF risk. X-axis represents the odds ratio of AF per one standard increase in the blood protein. **C**: Results of colocalization analysis on 27 blood proteins in relation to AF. High support (red) means *PH4* > 0.8 and otherwise *PH4* < 0.8 for blue. **D**: Cohort replication of the association between ADM protein and AF risk. X-axis represents the hazard ratio of AF per one standard increase in the blood protein. Model 1 was adjusted for batch effect, age, and sex; model 2 was adjusted for batch effect, age, sex, body mass index, education, baseline cardiovascular disease, smoking, alcohol intake, physical activity, and diet; and model 2 wa adjusted for all factors above plus levels of estimated glomerular filtration rate, lipids, glucose, and blood pressure.

### Modifiable factors and AF

Of the 26 studied modifiable exposures, 15 were associated with AF at the nominal significance level (**Figure 7A**). After multiple testing correction based on FDR, genetically proxied obesity, smoking liability, higher systolic and diastolic blood pressure, type 2 diabetes liability, lower high-density lipoprotein cholesterol levels, short sleep duration, and insomnia were associated with an increased risk of AF. The associations for obesity, smoking, and blood pressure remained after Bonferroni correction (**Table S14**). The associations also remained in sensitivity analyses (**Table S14**). Horizontal pleiotropy was detected for the association of genetic liability to type 2 diabetes with AF risk and genetically predicted low-density lipoprotein cholesterol levels with AF risk (**Table S14**).

**Figure 7.**
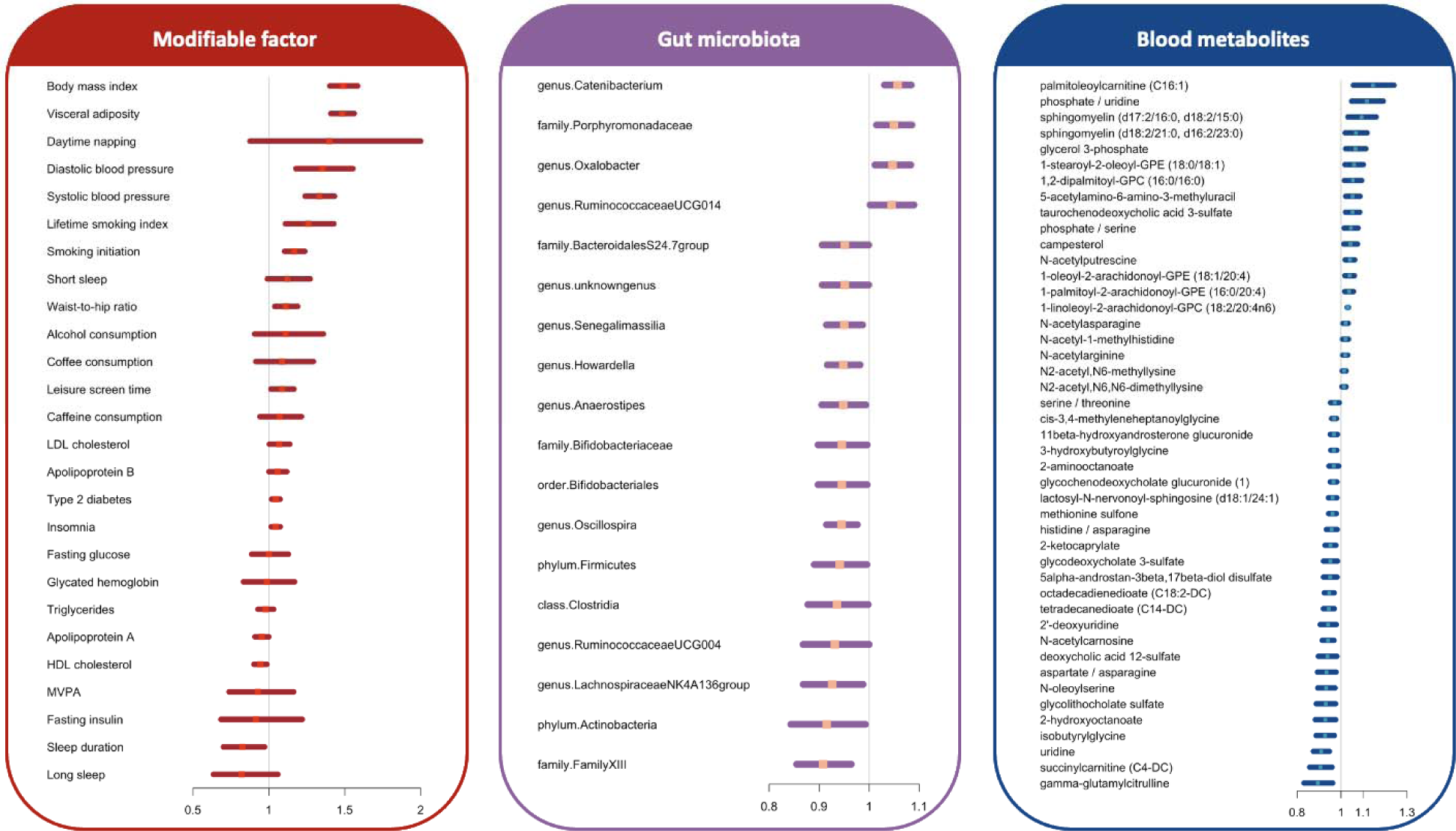
Associations of genetically proxied modifiable factors, gut microbiota, and blood metabolites and metabolite ratio with the risk of atrial fibrillation. Abbreviations: HDL, high-density lipoprotein; LDL, low-density lipoprotein; MVPA, moderate-to-vigorous physical activity. The x-axis indicates the odds ratio of AF. We showed associations between all studied modifiable factors and AF risk. For gut microbiota, the figure shows the associations with *P* value < 0.05. Given that many associations for blood metabolites were identified at the *P* value < 0.05, the figure shows associations with FDR < 0.05.

### Gut microbiota and AF

We examined the associations of 211 gut microbiotas with AF risk. Genetically predicted eighteen gut microbiotas were associated with AF at *P* < 0.0 5**Fi**(**gure 7B**). One association persisted after FDR or Bonferroni corrections (**Table S15**). Genetically predicted high abundance of the *genus.Catenibacterium.id.2153* was associated with an elevated risk of AF (**Table S16**).

### Blood metabolites and AF

Among 352 annotated metabolites and metabolite ratios, 45 were associated with AF after FDR corrections (**Figure 7C**), and 11 were identified using Bonferroni corrections (**Table S16**). These associations were consistent in sensitivity analyses, and we did not detect any indication of horizontal pleiotropy in the MR-Egger intercept test (**Table S16**).

## Discussion

In this study, we performed an updated GWAS meta-analysis of AF, including nearly 1.3 million individuals, and identified 215 loci, among which 91 were novel. Our study encompassed a series of in silico functional analyses spotlighted multiple candidate loci. Pleiotropy assessments revealed shared genetic etiologies between AF, cardiovascular comorbidities, and cardiometabolic traits. The PRS was a good predictor of AF risk when combined with age, sex, and basic cardiometabolic risk markers and correlated with multiple circulatory, endocrine, and respiratory-system comorbidities. Multiple omics-MR analyses uncovered modifiable factors, blood proteins, gut microbiota, and circulating metabolites with potentially causal roles in the development of AF. Findings on certain proteins, such as ADM, fibronectin fragment 3, and interleukin-6 receptor, may provide therapeutic hints.

Our updated GWAS confirmed all loci revealed in previously published large-scale GWASs^4–8^, including the strongest signal near *PITX2* gene. One rare and novel locus (i.e., rs532342679) near *NEDD1* was found to have a significant effect size on AF liability. This gene encodes the protein NEDD1 (neural precursor cell expressed developmentally down-regulated protein 1), a centrosomal protein essential in mitosis. NEDD1 protein is also involved in significant recruitment pathways of γ-TuRC (γ-tubulin ring complex) to the centrosome, which may influence embryonic development ^45^ and the functions of striated muscle cells (like, cardiomyocytes).^46^ Impaired reorganization of centrosome structure has been recently associated with infantile dilated cardiomyopathy. ^47^ *NEDD1* gene has also been revealed to be associated with body mass index and obstructive sleep apnoea, ^48^ which are risk factors for AF. Of note, the association for this locus was unavailable in the FinnGen study. However, variants near *NEDD1* gene were likely to be associated with AF risk, albeit not at the genome-wide threshold (*P* = 1.21×10^-4^ for rs34255398 or rs398039986).

In observational studies, AF has been associated with other cardiovascular comorbidities, such as CAD, HF, VTE, and stroke. ^1^ Our study supported the causality of these associations and the overall increased risk of circulatory diseases using the PRS-PheWAS analysis in the UK Biobank study. Our results of Genomic SEM analysis further provided genetic insights into the shared etiologies between AF and these cardiovascular comorbidities. For example, we found one locus (rs1537373 near *CDKN2B-AS1*) was shared by studied cardiovascular disease, except VTE. *CDKN2B* expression has been revealed to play a role in atherosclerosis^49^ by influencing postprandial triacylglycerol clearance ^50^ and impairing hypoxic neo-vessel maturation via impacting growth factor β signaling^51^. Another locus near *LPA* gene that determines the levels of lipoprotein(a) was found to be associated with the risk of AF, CAD, and HF, which is in line with previous findings.^52^

We also perfumed analyses to explore the shared genetic basis between AF and cardiometabolic traits and found many overlapping loci. Three AF-associated loci (rs10740129 near *JMJD1C*, rs2370982 near *NRXN3*, and rs9931494 near *FTO*) appeared to have universal effects on included cardiometabolic traits. The loci near *NRXN3* and *FTO* had concordant effects on AF and cardiometabolic phenotypes, which indicates that the alternations of cardiometabolic profile may be the molecular pathways linking the two loci and AF development. However, the locus near *JMJD1C* had opposite influences on AF and most cardiometabolic traits, except for low-density lipoprotein cholesterol. Although no underlying explanations, *JMJD1C* gene has been found to be involved in lipogenesis ^53^ and sex hormone regulation ^54^, which may affect AF risk^55^ independent of cardiometabolic profile.

AF is a chronic cardiovascular condition that may contribute to risk of stroke, heart failure, sudden death, or other complications needing hospitalization. ^56^ However, given that approximately 30% of AF patients are asymptomatic, early diagnosis of AF is still challenging, as apparent from many patients first being diagnosed after suffering a stroke.^56^ Electrocardiogram screening among the high-risk population seems promising. ^57^ However, no existing prediction scores for high-risk population identification have the potential for being widely adopted in the clinical setting.^58^ However, these scores did not consider genetic factors.^58^ In this study, the PRS score coupled with age, sex, and basic clinical features was found to be a good predictor of incident AF risk in Europeans, which may provide clues for the potential utilities of genetic information in AF high-risk population identification. Of note, this exploration is preliminary and further research is necessary to test the applicability and cost-effectiveness of this approach in a population-wide setting.

Our study using MR analysis identified several circulating proteins that associate with genetically predicted AF risk, which highlights potential therapeutic opportunities for drugs targeting these proteins, as well as insight into AF pathogenesis. Our MR analyses also identified several modifiable risk factors for AF, in particular obesity, high blood pressure, and cigarette smoking. These findings confirmed traditional epidemiological evidence^59^ and highlight the importance of reducing obesity, hypertension, and smoking in AF prevention. Gut microbiota and their bioactive metabolites gene rate health effects and have been linked to AF; however, which bacteria play a role in AF and the underlying mechanisms remain largely understood.^60^ Our current study found that genetically predicted higher abundance of *genus.Catenibacterium* was associated with an increased risk of AF. Even though this association was scarcely explored, the findings on *genus.Catenibacterium* in relation to cardiovascular risk have been conflicting. In a study among Tibetan Highlanders, *genus.Catenibacterium* were enriched in those suffering from CAD compared to healthy controls. ^61^ The abundance of this genus was also found to be higher among individuals with a healthier plant-centered diet that is related to lower risk of cardiovascular disease. ^62^ However, *genus.Catenibacterium* was found to be depleted among individuals with high versus low cardiovascular risk profile.^63^ More studies are needed to clarify the associations of gut microbiota, another potentially modifiable factor, with AF risk. Our study also identified several blood metabolites that may play role in AF development. These findings were generally consistent with previous results. For example, our findings on cis-3,4-methyleneheptanoylglycine supported the association between altered acylcarnitine metabolism and incident AF in the Malmö Diet and Cancer Study.^64^ In addition, our inverse association between uridine and AF was in line with the results in the Atherosclerosis Risk in Communities Study.^65^

There are many strengths of this study. First, we revealed many novel loci for AF using GWAS meta-analysis, including many cases (defined consistently across studies) and controls, and prioritized candidate genes from different angles. Second, based on known and novel genetic signals, we tested the utility of genetic and non-genetic factors in AF risk prediction and systematically explored AF-associated comorbidities. Third, we used varying methods to investigate the shared genetic etiological basis between AF, cardiovascular comorbidities, and cardiometabolic phenotypes. Fourth, we used different data and study designs to triangulate the associations of plasma proteins with AF and revealed potential therapeutic targets. Finally, we performed a wide-angle MR to gene rate evi dence to delineate pathological mechanisms underlying AF.

Limitations deserve to be discussed when interpreting our findings. First, our GWAS meta-analysis included only populations of European ancestry, which might restrict the generalizability of our results to other populations. Second, candidate prioritization and pathway analysis heavily relied on bioinformatics methods. These derived signals need confirmation using complementary approaches. Third, prospective data for protein-AF associations were available for a few proteins in SIMPLER cohorts. Whether the associations of other proteins with AF can be triangulated needs to be verified. Likewise, the same concern was raised up for the associations of gut microbiota and blood metabolites with AF. Fourth, we might have inadequate power in some analyses, such as for certain protein colocalization analyses.

Our study revealed novel loci genetic contributors to AF and shared genetic etiology between AF, cardiovascular comorbidities, and cardiometabolic traits. The AF-PRS, coupled with age, sex, and basic clinical features, showed a good prediction of the incidence AF risk. Omics-wide MR analysis revealed the underlying pathological complex of AF and potential therapeutic targets. Collectively, we provide translatable insights into AF risk prediction, pathophysiology and downstream sequelae.

## Acknowledgments

We want to acknowledge the participants and investigators of SIMPLER for provisioning of facilities and experimental support. SIMPLER receives funding through the Swedish Research Council under grant numbers 2017-00644, 2017-06100 and 2021-00160, and from Olle Engkvist Byggmästares stiftelse (SOEB). The computations were performed on resources provided by SNIC through Uppsala Multidisciplinary Center for Advanced Computational Science (UPPMAX) under Project simp2021005. We also want to acknowledge the participants and investigators of the UK Biobank study and the FinnGen study. This research was conducted using the UK Biobank study under application Number 66354.

## Author contributions

S.Y., Y.L, X.S. and S.C.L. conceived and designed the study. S.Y., Y.L., F.X., L.W., and X.L. undertook the statistical analyses. S.Y. and Y.L. wrote the first draft of the manuscript. All authors provided important comments to the manuscript and approved the final version of the manuscript.

## Data availability

The GWAS summary statistics for atrial fibrillation from Nielsen JB GWAS (http://csg.sph.umich.edu/willer/public/afib2018/), atrial fibrillation from the FinnGen study R8 (https://storage.googleapis.com/finngen-public-data-r8/summary_stats/finngen_R8_I9_AF.gz), coronary artery disease (https://www.ebi.ac.uk/gwas/publications/36474045), ischemic stroke (https://www.ebi.ac.uk/gwas/publications/36180795), heart failure (https://cvd.hugeamp.org/datasets.html), venous thromboembolism (https://www.decode.com/summarydata/), body mass index (https://www.ebi.ac.uk/gwas/publications/30239722), waist-to-hip ratio (https://www.ebi.ac.uk/gwas/publications/30239722), lipids (http://csg.sph.umich.edu/willer/public/glgc-lipids2021/results/ancestry_specific/), blood pressure (https://www.ebi.ac.uk/gwas/publications/30224653), type 2 diabetes (http://diagram-consortium.org/downloads.html), blood proteins (https://www.decode.com/summarydata/), and gut microbiota (https://www.mibiogen.org/) are publicly available. The summary statistics for this GWAS meta-analysis will be deposited at Zenodo when the paper is published.

## Code availability

Publicly available software was used to perform the analyses. Code is available from the corresponding author upon reasonable request.

