## Supplementary methods and figures for "Deciphering the genetic architecture of atrial fibrillation offers insights into disease prediction, pathophysiology and downstream sequelae"

### **1. Study participants, phenotyping, genotyping, and quality control**

**HUNT (The Nord-Trøndelag Health Study):** The HUNT survey, which began in 1984, is a population-based health study carried out in Nord-Trøndelag, Norway.^1^ To participate in the HUNT Study, informed consent is required, and the study has been approved by the Data Inspectorate and the Regional Ethics Committee for Medical Research in Norway. The genotyping was performed at the Norwegian University of Science and Technology (NTNU) using the Illumina HumanCore Exome v1.0 and v1.1, and quality control was conducted at both the marker and sample levels. Furthermore, variants from the HUNT low-pass genomes were imputed into The Haplotype Reference Consortium (HRC) samples using Minimac3, and only variants with imputation r^2^ values exceeding 0.3 were selected for analysis. The genetic associations with atrial fibrillation were estimated by a generalized mixed model including covariates birth year, sex, genotype batch, and principal components 1-4 as implemented in SAIGE.

**deCODE:** The study collected data on atrial fibrillation at Landspitali, The National University Hospital, in Reykjavik, and Akureyri Hospital (the two largest hospitals in Iceland) from 1987 to 2015. Controls were 358,161 Icelanders recruited through different genetic research projects at deCODE genetics, excluding those in the atrial fibrillation cohort. The study was approved by the Icelandic Data Protection Authority and the National Bioethics Committee of Iceland (no. VSNb2015030021). The research is centered on 15,220 Icelanders who took part in various disease projects at deCODE genetics, and whole-genome sequence data was used for the analysis. The sequencing process employed Illumina standard TruSeq methodology, and the mean depth was 35× (s.d. 8).^2^ The identification of autosomal SNPs and indels was done using Genome Analysis Toolkit version 3.4.0, with variants that failed quality control being removed following Genome Analysis Toolkit best practices. The SNPs and indels identified from sequencing were then imputed into 151,677 Icelanders and their close relatives who had been genotyped using Illumina SNP chips (familial imputation). Variants for the meta-analysis were selected based on matching with either the 1000 Genomes Project reference panel (Phase 3) or the Haplotype Consortium reference panel, taking into account factors such as allele, frequency, and correlation matching. To test for the association between SNPs and atrial fibrillation, logistic regression was used. Additionally, LD score regression was employed to adjust for inflation in test statistics due to cryptic relatedness and stratification.

**MGI (The Michigan Genomics Initiative):** The MGI cohort is a hospital-based study conducted at Michigan Medicine in the United States, which was approved by the Institutional Review Board of the University of Michigan Medical School. Genotyping was carried out at the University of Michigan using the Illumina Human Core Exome v1.0 and v1.1, followed by quality control at the sample and marker level. Variants from the Haplotype Reference Consortium (HRC) reference panel were imputed using the Michigan Imputation Server (refer to URLs), and variants with imputation r^2^ >0.3 were retained for further analysis. The association between the variants and atrial fibrillation was assessed using the Firth bias-corrected logistic likelihood ratio test.

**DiscovEHR:** The ongoing MyCode Community Health Initiative of the Geisinger Health System, USA contributed to the DiscovEHR collaboration cohort, which is a hospital-based cohort consisting of 58,124 genotyped individuals of European ancestry.^3^ The Geisinger Institutional Review Board approved the study. Genotyping was conducted on the Human OmniExpress Exome Beadchip at Illumina after aliquots of DNA were sent. Principal component analysis was used to identify individuals of European ancestry, and imputation to the HRC reference panel was performed using the Michigan Imputation Server. Variants with imputation r^2^ >0.3 and MAF >0.001 were retained for analysis. The BGEN dosage files were analyzed using BOLT-LMM, and variants were tested for their association with atrial fibrillation under an additive genetic model.

**The UK Biobank study:** The UK Biobank is a cohort based on the general population that was collected from multiple locations throughout the United Kingdom. The study received approval from the North West-Haydock Research Ethics Committee (REC reference: 21/NW/0157), and all participants provided electronic consent. Participants were genotyped using two similar genotyping arrays designed specifically for the UK Biobank (Applied Biosystems UK BiLEVE Axiom Array and UK BioBank Axiom Array). Phasing and imputation were performed by the UK Biobank analysis team, utilizing the HRC reference panel and UK10K haplotype resource. The study focused on HRC-imputed markers, and analysis of genetic associations with atrial fibrillation was conducted among people of white British ancestry, utilizing a generalized mixed model implemented in SAIGE.

**AFGen consortium (The Atrial Fibrillation Genetics consortium):** The AFGen consortium is composed of 33 studies, mainly consisting of participants with European ancestry.^4^ The identification of atrial fibrillation cases involved documented atrial fibrillation on an electrocardiogram or one in-patient or two out-patient diagnoses of atrial fibrillation, while referents were free of atrial fibrillation. Ethics committees or institutional review boards approved the study, and informed consent was obtained from all cases and referents. Each study conducted genotyping and imputation using the 1000 Genomes Project Phase 1 reference panel, with pre- and post-GWAS filtering steps performed for quality control. The meta-analysis of GWAS results was conducted using an inverse-variance-weighted fixed-effects model and the METAL software.

**The FinnGen study (R8):** The FinnGen research project is a collaboration between public and private entities that integrates genotype data from Finnish biobanks with digital health record data from Finnish health registries. Genotyping was performed using Illumina (Illumina Inc., San Diego, CA, USA) and Affymetrix arrays (Thermo Fisher Scientific, Santa Clara, CA, USA), with genotype calls made using GenCall and zCall algorithms for Illumina and AxiomGT1 algorithm for Affymetrix data. Sample-wise quality control excluded individuals with ambiguous gender, high genotype missingness (>5%), excess heterozygosity (+-4SD), and non-Finnish ancestry. Variant-wise quality control excluded variants with high missingness (>2%), low Hardy-Weinberg equilibrium P-value (<1e-6), and low minor allele count (MAC<3). Imputation of genotypes was performed using the population-specific SISu v4.0 reference panel. The genetic associations were estimated using Plink. Detailed information can be found: <https://finngen.gitbook.io/documentation/>

**SIMPLER cohorts:** SIMPLER stands for Swedish Infrastructure for Medical Population-Based Life-Course and Environmental Research, which includes two Swedish cohorts that are the Swedish Mammography Cohort (SMC, initiated in 1987) and the Cohort of Swedish Men (COSM, initiated in 1997). The clinical examinations in sub-cohorts of SMC (SMCC) and COSM (COSMC) were conducted in 2003-2009 and 2010-2019, respectively, which lead to a total of almost 13 500 participants in the two sub-cohorts. In the whole SIMPLER, there were approximately 37 000 participants with genetic data sequenced by Infinium Global Screening Array (Illumina). Proteome profile has been analyzed using the panels CVD II, CVD III and metabolism from Olink Proteomics using Li/Hep blood plasma. The proteome profile covers the clinical participants, approximately 13,500 participants. The AF diagnostic data were extracted from from the Swedish National Patient Register, which covers nearly all hospital-based inpatient and outpatient care. In COSMC, staff at Eurofins Genomics (Ebersberg, Germany) extracted DNA from 4 ml EDTA whole blood with use of QIAamp DNA Blood Midi Kit” (Cat. No. 51185) from Qiagen (Hilden, Germany). Subsequently, samples were genotyped at Eurofins Genomics with the Illumina Infinium Global Screening Array version 3 (GSAv3; Illumina, San Diego, CA, USA). Sample exclusion filters applied were: (1) samples with discordant sex information when comparing reported sex and sex determined by the X-chromosome; (2) non-European ancestry; (3) heterozygosity outliers -/+3*IQR from Q1/Q3; (4) low sample call rate (<98 %); (5) HWE exact *p*-value <1 × 10−7 (--hwe midp); (6) minor allele count <20; (7) markers not present in 1000G/HRC with matching alleles; (8) allele frequency difference > 0.15 compared with 1000G/HRC. We imputed data by use of chr1-22,X: Michigan Imputation Server v1.2.4 using Eagle v2.4 + minimac v4 and both 1000G phase3 (v5) in tgp.ph 3/ and HRC v1.1 in in hrc1.1/ as reference panels. The final genetic dataset included approximately 7.8 million markers. In SMCC, staff at the Biobank at Karolinska Institutet extracted DNA from 400 μl EDTA whole blood with the Chemagen STAR DNA Blood 400 kit (Perkin Elmer, Waltham, MA, USA) using a ChemagicStar-robot (Hamilton, Reno, NV, USA) based on magnetic bead separation. Subsequently, samples were genotyped at the SNP&SEQ Technology Platform, Science for Life Laboratory, Uppsala University with the Illumina Infinium Global Screening Array Multiple Disease version 1 (GSAv1; Illumina, San Diego, CA, USA). We used the same exclusion filters for quality control. For imputation of chr1-22 the Michigan Imputation Server v1.0.4 using Eagle v2.3 + minimac v3 was used and for chrX the Michigan Imputation Server v1.2.4 using Eagle v2.4 + minimac v4, with reference panels 1000G phase3 (v5) in tgp.ph 3/ and HRC v1.1 in in hrc1.1/. The final genetic dataset included approximately 7.8 million markers. The genetic associations with atrial fibrillation were estimated by a generalized mixed model as implemented in SAIGE.

### **2. Transcriptome-wide association study (TWAS)**

We utilized S-MultiXcan to integrate GTEx v8 gene expression and splicing data with summary statistics from our AF GWAS meta-analysis to identify genes associated with atrial fibrillation.^5,6^ The strength of S-MultiXcan lies in its ability to harness the significant overlap of eQTLs across various tissues and contexts, enhancing the capacity to pinpoint potential target genes. Furthermore, S-MultiXcan incorporates evidence from multiple panels via multivariate regression, which naturally considers the inherent correlation structure. This method enables us to explore the phenotypic consequences of tissue-specific gene expression variation inferred from GWAS summary statistics. We performed MASHR-based models which are the pre-trained models for gene expression, and transcript splicing were acquired from <http://predictdb.org/>. Bonferroni adjustment was performed to account for multiple testing (22,535 gene–tissue pairs for eQTL TWAS; 139,035 splicing event–tissue pairs for sQTL TWAS).

### **3. Multi-trait GWAS (Genomic-SEM analysis)**

Multivariate models were implemented using the GenomicSEM package in R.^7^ We applied a common factor model. Detailed description of the GenomicSEM package can be found at <https://github.com/GenomicSEM/GenomicSEM>. The method begins by fist estimating the genetic covariance matrix using GWAS summary statistics and a multivariate extension of LDSC. Then each model is specified using a system of equations. Finally, the parameter(s) of interest are regressed on each SNP. A common factor model specifies a latent variable which represents the shared variance among related traits. This latent trait can variably influence each of the downstream traits (in this case AF and included other cardiovascular diseases). A common factor GWAS thus considers the effects of genetic variants on the shared heritability of related traits. Here, the common factor GWAS considers a latent trait which influences AF and other cardiovascular diseases. Here, we specified a system of equations where 1) our target phenotype of interest (AF) was regressed on each SNP, and 2) the supporting phenotypes (in this case the cardiac MRI traits) were regressed on the target phenotype (other cardiovascular diseases). The results of this GWAS represent more precise SNP-effects on AF, and by virtue of increasing precision also improves power for novel discovery.

### **4. Mendelian randomization analysis**

Employing genetic variants as instrumental variables for an exposure, Mendelian randomization (MR) analysis is an epidemiological approach that can reinforce causal inference in an exposure-outcome association using observational genetic data.^8^ The approach can minimize residual confounding because genetic variants are randomly allocated at conception and thus generally unrelated to confounders, such as environmental and self-adopted factors. The random allocation of effect allele in MR design resembles the randomization process in randomized controlled trials. In addition, the method can diminish reverse causation because genetic variants used to proxy the effect of the exposure cannot be modified by the onset and progression of the outcome.

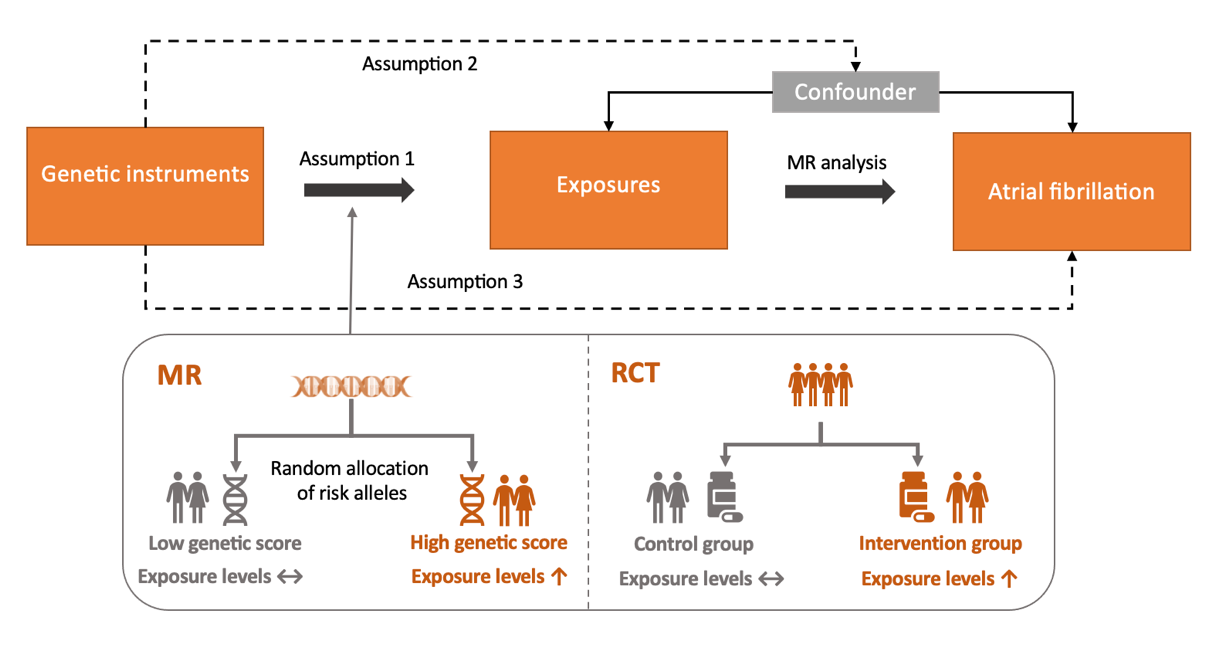

Figure. Directed acyclic graph (DAG) for Mendelian randomization analysis on atrial fibrillation. There are 3 important assumptions of MR analysis. The first assumption is that the genetic variants proposed as instrumental variables should be robustly associated with the exposure; the second assumption indicates that the used genetic variants should not be associated with any confounders; and the third assumption is that the selected genetic variants should affect the risk of the outcome merely through the risk factor, not via alternative pathways.

The inverse-variance weighted (IVW) method was used as the main statistical model.^9^ For traits with SNPs ≤ 3, we used the inverse variance weighted method under the fixed effects model to estimate the MR association with AF. Otherwise, the inverse variance weighted method under the multiplicative random effects was used. For traits with SNPs ≥ 3, we performed two sensitivity analyses that are the weighted median and MR-Egger regression methods to test the consistency of the results. Assuming that at least 50% of the SNPs are valid, the weighted median method can generate consistent causal estimates.^10^ The MR-Egger regression can detect and correct for possible pleiotropy, and the *P* value of the intercept > 0.05 indicates no horizontal pleiotropic effects.^11^ All analyses were 2-sided and performed using the TwoSampleMR^12^ package in R software (version 4.0.2).

### **5. Colocalization analysis**

We conducted colocalization analysis using the coloc R package^13^ to test whether identified associations between proteins and AF were driven by linkage disequilibrium. For each locus, the Bayesian method assessed the support for the following five exclusive hypotheses: 1) no association with either trait; 2) association with trait 1 only; 3) association with trait 2 only; 4) both traits are associated, but distinct causal variants were for two traits; and 5) both traits are associated, and the same shares causal variant for both traits. The analysis provides posterior probabilities for each hypothesis testing (H0, H1, H2, H3, and H4). We set prior probabilities of the SNP being associated with trait 1 only (p1) at 1 × 10^−4^; the probability of the SNP being associated with trait 2 only (p2) at 1 × 10^−4^; and the probability of the SNP being associated with both traits (p12) at 1 × 10^−5^ two signals were considered to have strong evidence of colocalization if the posterior probability for shared causal variants (PH4) was ≥0.8.

The colocalization analysis was based on summary-level statistics of genetic associations with levels of 4907 circulating proteins from a large-scale protein quantitative trait loci (pQTL) study in 35,559 Icelanders.^14^ Proteomic profiling was performed by a multiplexed, modified aptamer-based binding assay (SOMAscan version 4). The levels of protein were rank-inverse normal transformed by age and sex. The residuals were standardized using rank-inverse normal transformation and the standardized values were treated as phenotypes in the genome-wide association analyses under the BOLT-LMM linear mixed model.

### **6. Cohort analysis on the association between ADM protein and incident AF in SIMPLER**

We conducted a prospective cohort analysis in SIMPLER to replicate certain MR associations for blood proteins that were measured using the Olink platform. This analysis was based on the clinical sub-cohorts of the Swedish Infrastructure for Medical Population-based Life-course and Environmental Research (SIMPLER) that includes the Swedish Mammography Cohort (SMC) and Cohort of Swedish Men (COSM). The clinical examinations in sub-cohorts of SMC and COSM were conducted in 2003–2009 and 2010–2019, respectively. Samples of fasting blood were collected at health examinations and participants were asked to fill in questionnaires on diet, health and lifestyle. After removing 1518 individuals with baseline AF from 12,314 participants with available protein data, we included 10,796 participants from the two sub-cohorts in the analysis. Detailed information on two sub-cohorts and questionnaires can be found on the SIMPLER website (<https://www.simpler4health.se/>).

Venous blood samples were collected after a 12-h overnight fast and immediately centrifuged and stored at −80°C until analysis. In total, 276 protein biomarkers were analyzed using three high-throughput multiplex immunoassays: the Olink Proseek Multiplex CVD II, CVD III and Metabolism (Olink Bioscience). Each assay measured 92 selected cardiovascular disease- or metabolism-related proteins. The platform provides normalized protein expression values on a log2 scale standardized per analysis plate. Values below the limit of detection (LOD) were provided by the manufacturer and used as a protein selection criterium. The analyses were performed at SciLifeLab, Uppsala University, Sweden. We excluded proteins with more than 50% samples below the LOD. A small portion of specimens was set to missing given the analyzed sample did not pass the manufacturer's quality control (3.6%, 0.8% and 0.7% for CVD II, CVD III and metabolism panels, respectively). The analysis included 257 proteins.

AF cases were ascertained by a medical diagnosis of AF, either as the primary or contributing causes with diagnostic information (codes of International Classification of Diseases-9 and -10) from the Swedish National Patient Register, which covers nearly all hospital-based inpatient and outpatient care. Dates of death were obtained from the Swedish Death Registry. Individuals were followed up from the baseline until the date of diagnosis of AF, date of death, or end of follow-up (i.e., 31 December 2019), whichever came first.

Information on age, sex, education attainment, smoking, alcohol consumption, physical activity, and diet quality was obtained from self-administrated questionnaires. Diet quality was assessed by a modified Dietary Approaches to Stop Hypertension (mDASH) score.^15^ Body mass index (BMI, weight/height squared), estimated glomerular filtration rate (eGFR), levels of blood lipids and glucose, and blood pressure were measured in the health exam. We obtained data on baseline diagnosis of cardiovascular disease including coronary artery disease, heart failure, and stroke from Swedish National Patient Register.

Cox proportional hazards regression model with age as the underlying time scale was used to estimate the hazard ratios (HRs) and corresponding 95% confidence intervals (CIs) of the association between circulating proteins and incident AF risk. The assumption of proportionality was met as indicated by Schoenfeld residuals. Three models were used: (a) Model 1 adjusted for sex and batch; (b) Model 2 adjusted for sex, batch, BMI, educational attainment, baseline cardiovascular disease, smoking status, alcohol consumption, physical activity, and mDASH score; and (c) Model 3 adjusted for sex, batch, BMI, educational attainment, baseline cardiovascular disease, smoking status, alcohol consumption, physical activity, mDASH score, eGFR, low- and high-density lipoprotein cholesterol, triglycerides, blood pressure, and blood glucose levels.

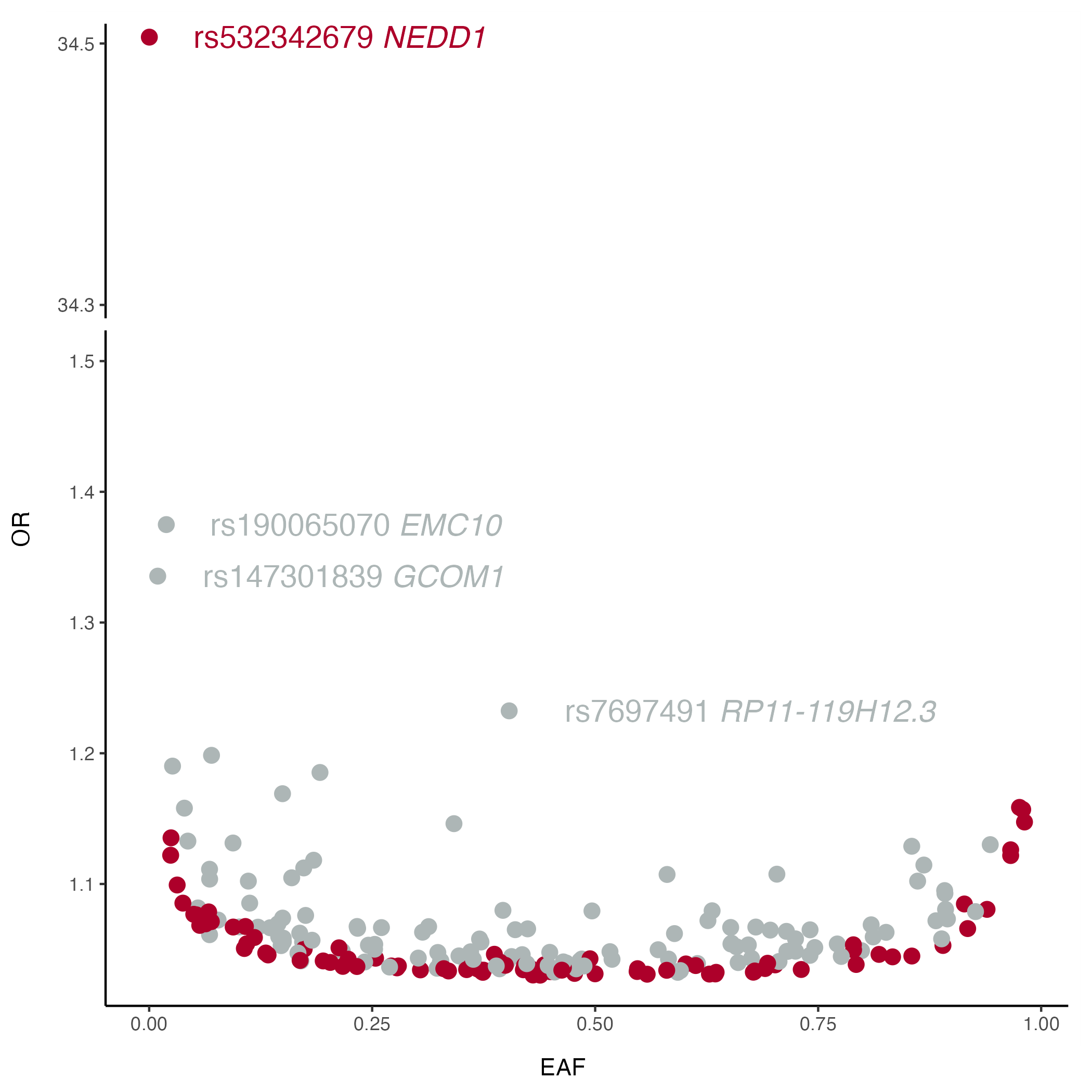

### **Figure S1. Effect size distribution.**

Odds ratio (OR) of AF for 215 lead variants in the GWAS meta-analysis against risk allele frequency. Variants with OR > 1.3 are annotated. The color of each point indicates whether the locus is known (gray) or novel (red).

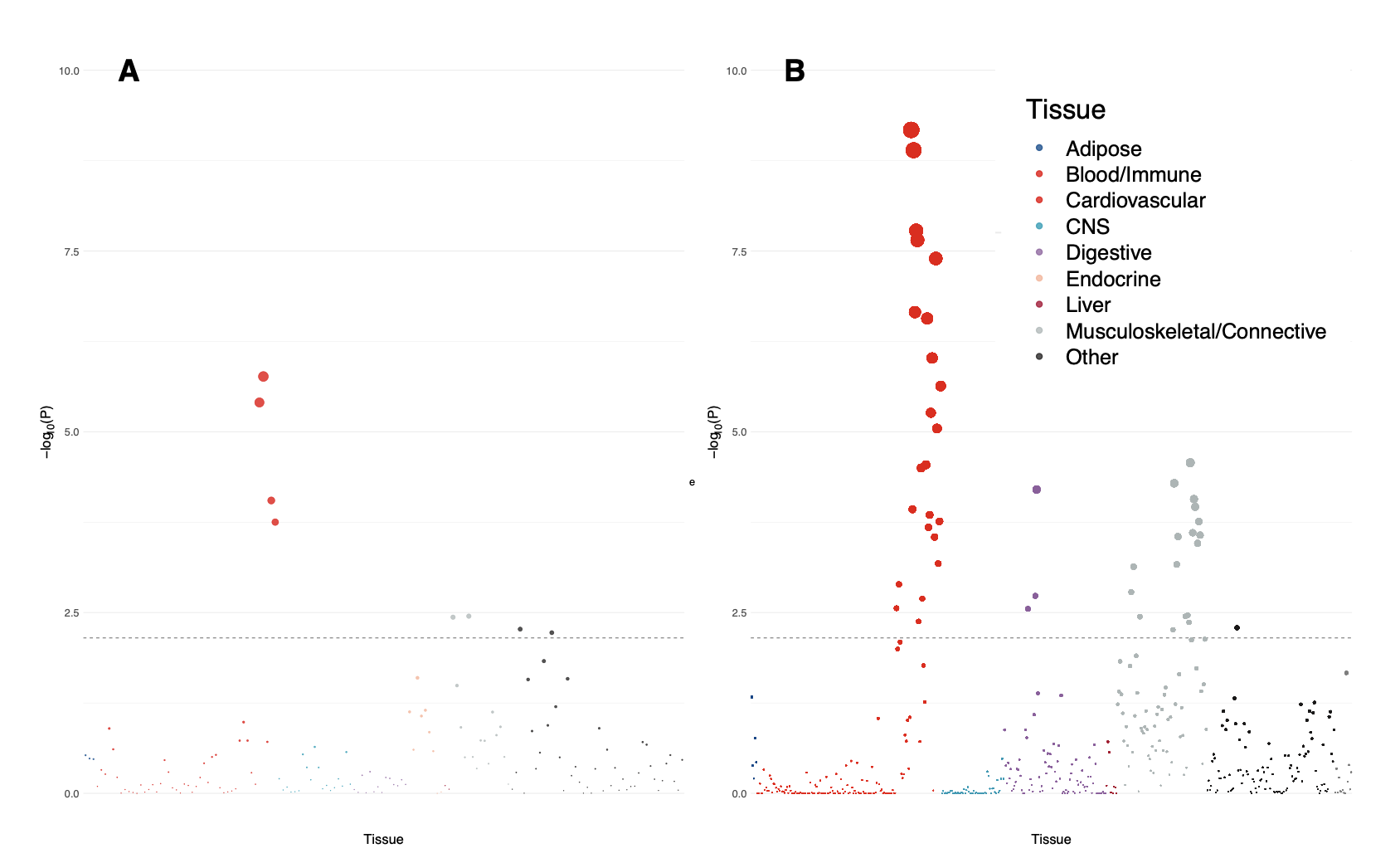

### **Figure S2. Tissue-specific enrichment using LDSC-SEG.**

LDSC-SEG was performed to identify tissue specific associations with AF. A) Associations between AF-associated loci and tissue-specific gene expression; and B) Associations between AF-associated loci and tissue-specific chromatin marks.

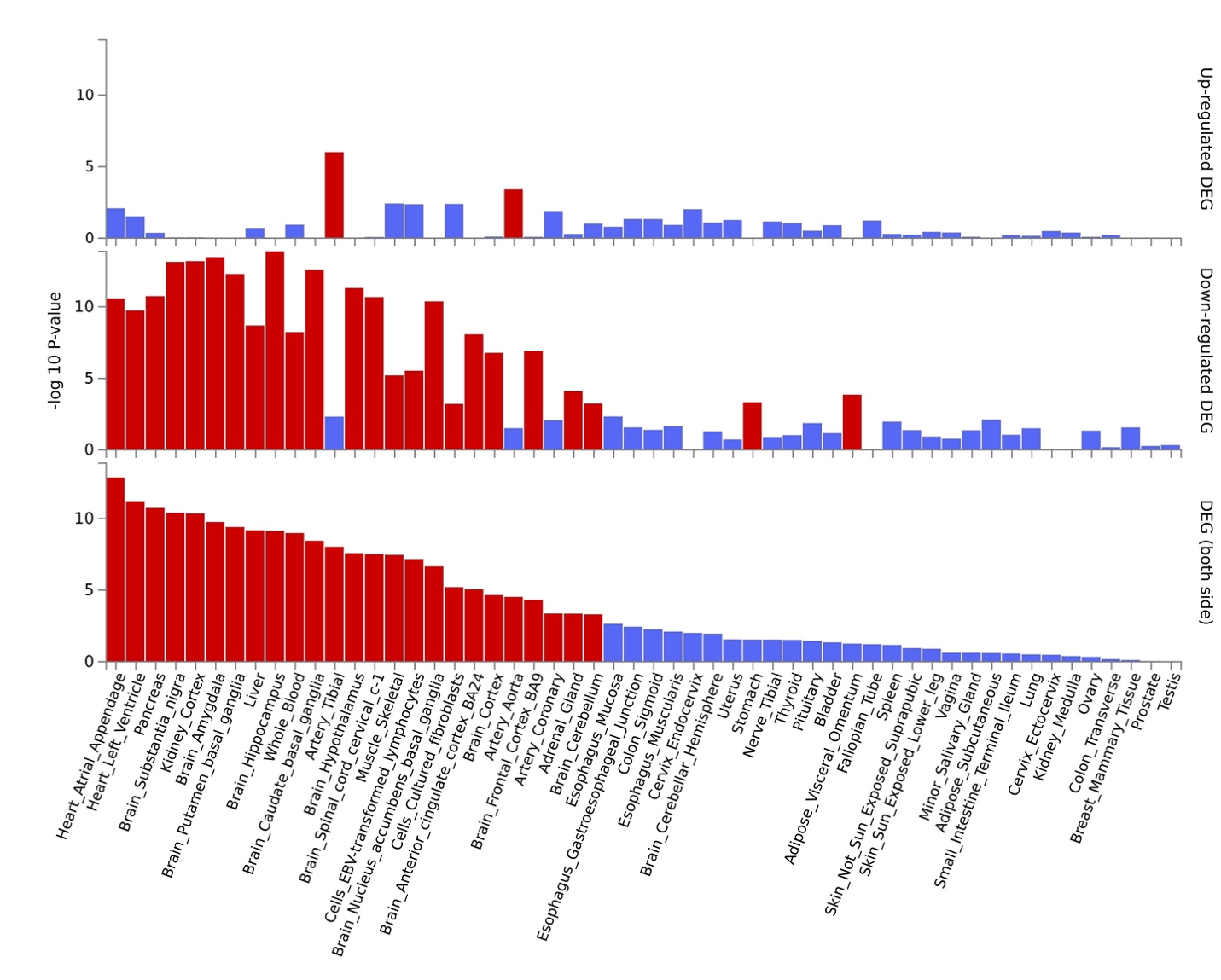

### **Figure S3. Tissue-specific enrichment using FUMA.**

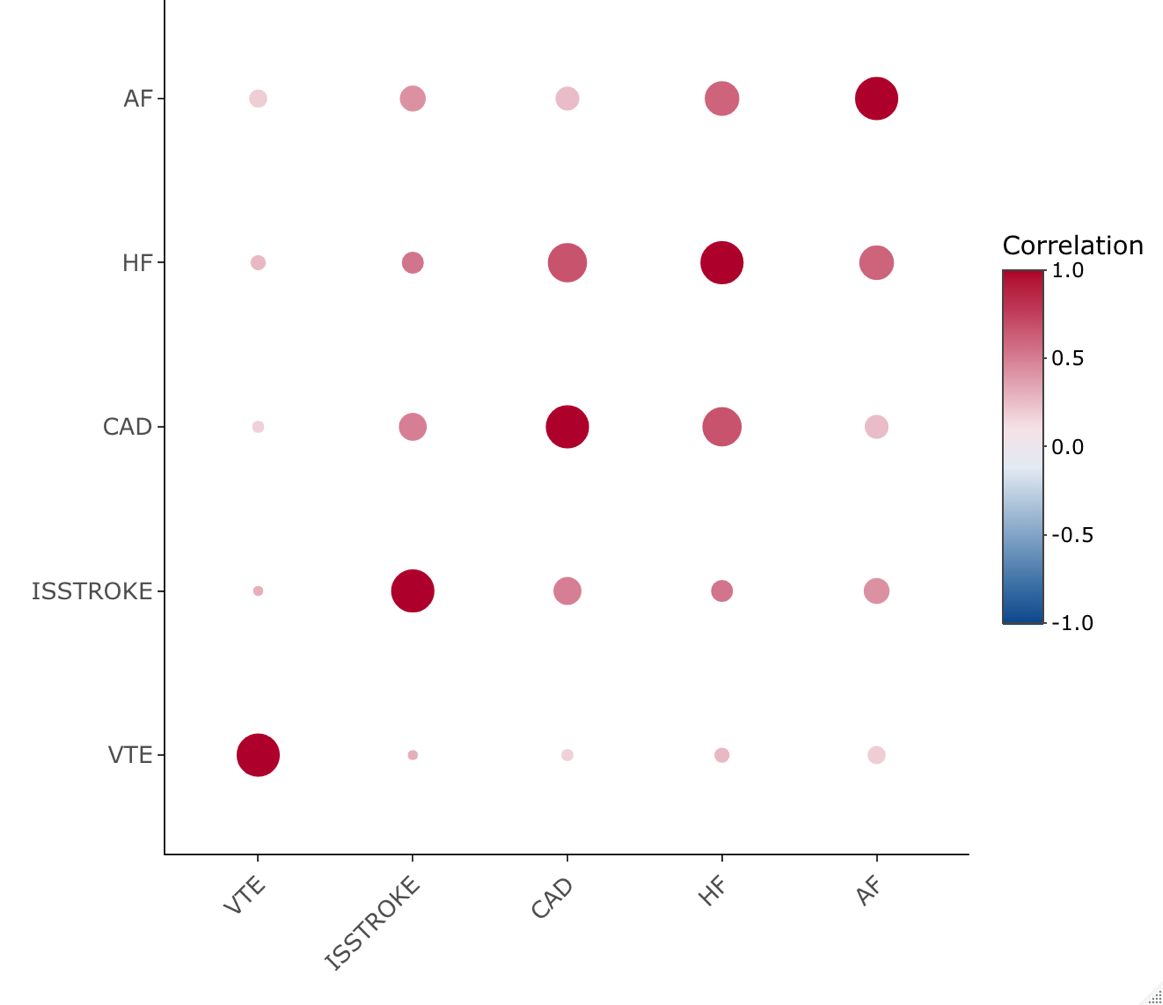

### **Figure S4. Genetic correlations among atrial fibrillation (AF) and other four major cardiovascular diseases.**

CAD, coronary artery disease; HF, heart failure; ISSTROKE, ischemic stroke; VTE, venous thromboembolism. The associations were with the *P* value < 7.13×10^-11^.

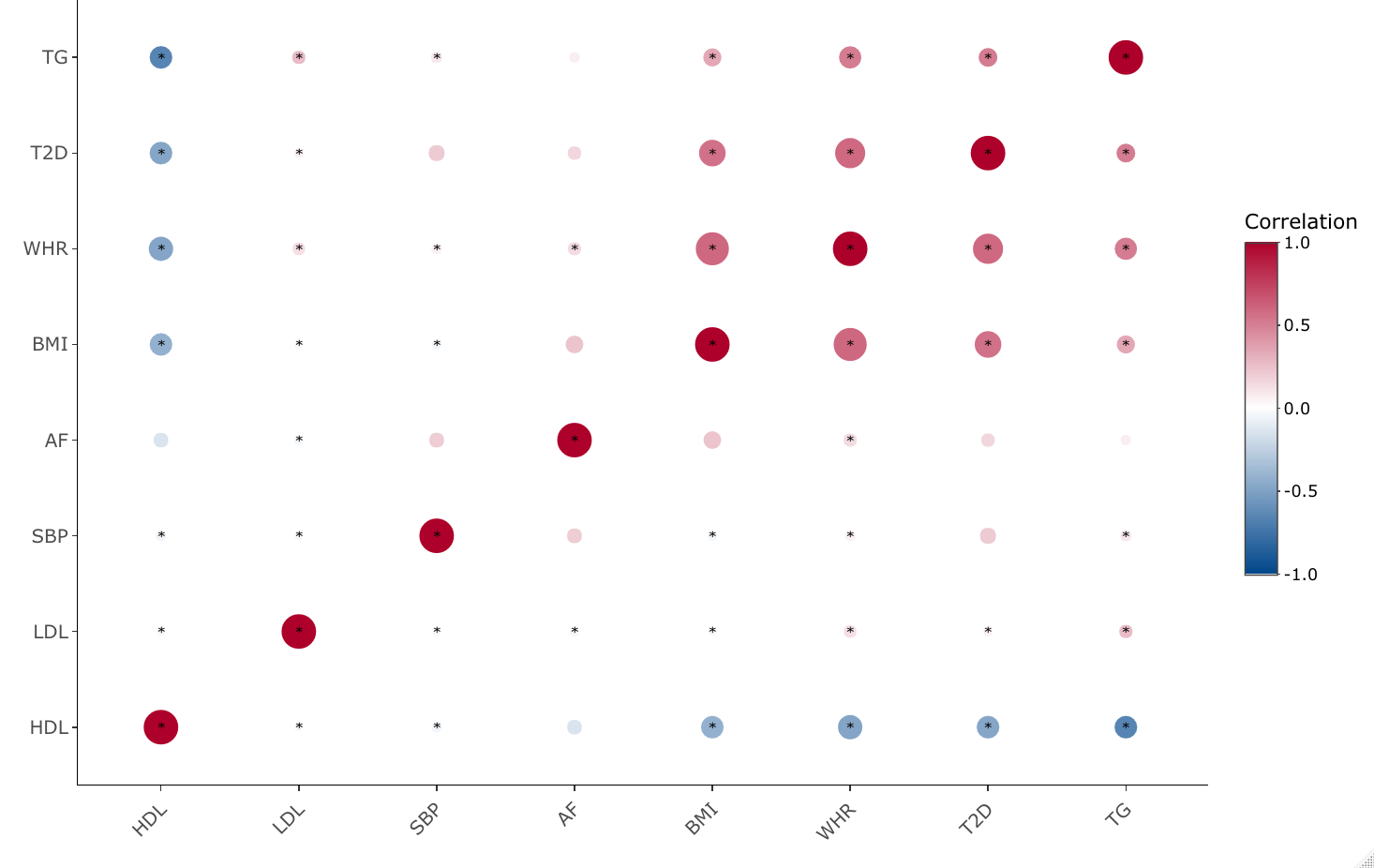

### **Figure S5. Genetic correlations among atrial fibrillation (AF) and 7 cardiometabolic traits.**

AF, atrial fibrillation; BMI, body mass index; HDL, high-density lipoprotein; LDL, low-density lipoprotein; SBP, systolic blood pressure; T2D, type 2 diabetes; TG, triglycerides; WHR, waist-to-hip ratio. Star mark indicates significant associations after Bonferroni correction (*P* < 0.002).
